# Serum cytokine and inflammatory markers in individuals with heroin use disorder: potential biomarkers for diagnosis and disease severity

**DOI:** 10.1101/2024.04.29.24306559

**Authors:** Eduardo R. Butelman, Yuefeng Huang, Flurin Cathomas, Pierre-Olivier Gaudreault, Panos Roussos, Scott J. Russo, Rita Z. Goldstein, Nelly Alia-Klein

## Abstract

Opioid use disorders cause major morbidity and mortality, and there is a pressing need for novel mechanistic targets and biomarkers for diagnosis and prognosis. Exposure to mu-opioid receptor (MOR) agonists causes changes in cytokine and inflammatory protein networks in peripheral blood, and also in brain glia and neurons. Individuals with heroin use disorder (iHUD) show dysregulated levels of several cytokines in blood. However, there is limited data on a comprehensive panel of such markers in iHUD versus healthy controls (HC), especially as a multi-target biomarker. We used a validated proximity extension assay for relative quantification of 92 cytokines and inflammatory proteins in serum of iHUD on medication assisted therapy (MAT; n=21), versus HC (n=24). Twenty-nine targets showed significant group differences (primarily iHUD>HC), surviving multiple comparison correction (p=0.05). This included 19 members of canonical cytokine families, including specific chemokines, interleukins, growth factors, and tumor necrosis factor (TNF)-related proteins. For dimensionality reduction, data from these 19 cytokines were entered into a principal component (PC) analysis, and PC1 scores were iHUD>HC (p<0.0001). A receiver-operating characteristic (ROC) curve analysis yielded an AUROC=91.7% (p<0.0001). This PC1 score remained a positive predictor of being in the HUD group in a multivariable logistic regression, which included demographic/clinical variables. Overall, this study shows a panel of cytokines that differ significantly between iHUD and HC, and provides a multi-target “cytokine biomarker score” for potential diagnostic purposes, and examination of disease severity.

## Introduction

Heroin use disorders (HUD) and other opioid use disorders pose significant challenges to society, resulting in substantial morbidity and mortality ^1^, including a 2022 provisional estimate from the CDC of approximately 83,000 deaths attributed to opioid-related causes ^2^. Heroin and other opioid compounds such as fentanyl act as agonists at mu-opioid receptors (MOR), which mediate the direct effects as well as long-term pathophysiology of these compounds, both in the periphery and central nervous system. Although effective medication-assisted therapies (MAT) exist for HUD, such as oral maintenance with the MOR agonist methadone or the partial MOR agonist/kappa-opioid receptor (KOR) antagonist buprenorphine, a significant proportion of individuals with HUD (iHUD) discontinue treatment or relapse ^3–5^. The underlying mechanisms for these undesirable outcomes remain unclear, highlighting an urgent need for novel mechanistically-based treatments. There is also a pressing need for objective and quantitative biomarkers for HUD ^6–10^. Due to their relative non-invasiveness and practicality, measurement of blood-based biomarkers have emerged as powerful approaches in the study of diverse neuropsychiatric disorders ^11,12^.

In addition to their actions on neuronal functions ^13,14^, MOR agonists and their cognate receptors interact with complex networks of cytokines (e.g., chemokines, interleukins, growth factors and tumor necrosis factor - related proteins) ^15,16^, signaling proteins that operate as interactive networks both in the periphery (e.g., in circulating leukocytes), and in central glia and neurons ^17,18^. Recent studies in preclinical models show that neuronal-glial interactions, often mediated by cytokines, are crucial in homeostatic functions including neuroplasticity and behavioral outcomes ^18–20^. Importantly, circulating leukocytes and the cytokines they release may mediate some of the neurobiological and behavioral consequences of HUD ^21,22^. Prior studies have detected differences in levels of some cytokines in iHUD compared to healthy controls (HC), with some discrepancies across studies, possibly due to methodological differences ^23–25^. However, there remains a scarcity of data using a comprehensive panel of cytokines from major cytokine families, including chemokines, interleukins, growth factors, and TNF-related proteins, in the context of clinical HUD ^7,21^.

The goals of this study were therefore twofold: First, to examine differences between iHUD and HC using a large and representative panel of cytokines and other inflammatory proteins ^11,26^, aiming to identify novel targets that are potentially related to the severity of HUD, including the trajectory of heroin exposure at an earlier age ^1,27^; these can be explored with variables such as age of onset of first and regular heroin use, as well as years of regular heroin use. Secondly, to develop an overall “cytokine biomarker score” via dimensionality reduction with principal component analysis (PCA), and determine if it can robustly differentiate iHUD from HC, taking into account major variables that may affect cytokine targets, especially age, body-mass index, sleep, and perceived stress ^28–32^.

## Methods

### Participants and Diagnostic procedures

Twenty-one iHUD and 24 age- and sex-matched HC were recruited for the current study. All iHUD were recruited from an inpatient drug addiction rehabilitation organization (Samaritan Daytop Village, NY). The HC were recruited from the surrounding communities, for matching purposes. This study was approved by the Institutional Review Board of the Icahn School of Medicine at Mount Sinai, and all participants provided written informed consent. All participants underwent a comprehensive clinical diagnostic interview, conducted by trained research staff under a clinical psychologist’s supervision, including the Addiction Severity Index (ASI) ^33^ and the M.I.N.I. neuropsychiatric interview ^34^ for DSM-5 diagnoses. **Inclusion criteria for all participants:** Ability to understand and give informed consent in English, and 18-65 years of age. **Inclusion criteria for iHUD specifically:** Meet DSM-5 criteria for opioid use disorder, with heroin as the primary drug of choice or reason for treatment. We did not exclude iHUD with DSM-5 diagnosis of a drug use disorder other than opiates, as long as heroin was the primary drug of choice and reason for seeking treatment, as iHUDs commonly use other drugs. All iHUD were inpatients in MAT and stabilized on methadone (n=17) or buprenorphine (n=4). **Exclusion criteria for all participants:** 1). DSM-5 diagnoses for psychotic disorders (e.g. schizophrenia) or neurodevelopmental disorders (e.g. autism). 2).

History of head trauma with loss of consciousness (>30 min). 3). Neurological disease of central origin, including seizures. 4). Cardiovascular disease including high blood pressure. 5). Active infectious diseases such as hepatitis B/C or HIV/AIDS. 6). Other active medical conditions, including metabolic, endocrinological, oncological, or autoimmune diseases. **Exclusion criteria HC specifically:** History of drug or alcohol use disorders or any psychiatric diagnoses.

### Demographic, behavioral, and clinical variables

In addition to sex, racial background, and age, BMI and hours of sleep in the night prior to testing were examined. Stress exposure, depression, and anxiety have been associated with changes in cytokine levels ^11,28,35^; we therefore examined perceived stress with the PSS-10 scale ^36,37^, self-rated anhedonia, dysphoria, pessimism, and fatigue with the Beck Depression Inventory (BDI-II) ^38,39^, and somatic and cognitive symptoms of anxiety with the Beck Anxiety Inventory ^40^. In the iHUD, we further examined methadone dose (documented report, or self-report if the former was unavailable) and duration of current abstinence from heroin, as well as the age of first heroin use, the age of regular use, and the number of years of regular use (excluding periods of abstinence) ^41,42^. A summary of demographic and clinical variables are in Table 1, and compared across groups with Mann-Whitney tests or Fisher’s exact tests.

**Table 1.** Demographics.

| **Variable** | **iHUD**<br>**(n=21)** | **HC**<br>**(n=24)** | **Group differences tests;**<br>**Mann-Whitney U or**<br>**Fisher's exact test** |
| --- | --- | --- | --- |
| **Age: mean (95%CI)** | 42.0 (38.5-45.5) | 40.7 (36.0-45.4) | NS |
| **Sex: M / F** | 17M / 4F | 15M / 9F | Fisher's test; NS |
| **Race: White / Black / Other** | 18 / 0 / 3 | 12 / 8 / 4 | Fisher's test; p = 0.006 |
| **BMI: mean (95%CI)** | 32.3 (29.4-35.3) | 26.7 (24.9-28.6) | U=110; p=0.0009 |
| **Sleep hours in the prior night:**<br>**mean (95%CI)** | 6.54 (5.6-7.5) | 6.51 (5.9-7.2) | NS |
| **PSS-10 Scores: mean (95%CI)\*** | 20.5 (17.7-23.3)<br>n=20 | 12.3 (9.5-16.0) | U=104; p=0.001 |
| **BDI-II Depression: mean (95%CI)\*** | 15.5 (9.9-21.2)<br>n=19 | 3.8 (1.8-5.7) | U=78; p=0.0001 |
| **BAI Anxiety: mean (95%CI)\*** | 14.1 (8.8-19.4)<br>n=20 | 2.8 (1.1-4.4) | U=85.5; p=0.0001 |
| **Age of onset of first use of heroin:**<br>**mean (95%CI)** | 23.6 (17.4-27.8)<br>n=20 | N/A | N/A |
| **Age of onset of regular use of heroin:**<br>**mean (95%CI)** | 25.1 (21.2-28.9)<br>n=20 | N/A | N/A |
| **Years of regular heroin use:**<br>**mean (95%CI)** | 8.5 (5.9-11.2)<br>n=19 | N/A | N/A |
| **Heroin abstinence duration in days:**<br>**mean (95%CI)** | 198 (130-268) | N/A | N/A |
| **Methadone daily dose in mg:**<br>**mean (95%CI)** | 91.9 (63-121)<br>n=17 | N/A | N/A |

Unless otherwise stated, n=21 for iHUD (of which n=17 with methadone and n=4 with buprenorphine MAT), and n=24 for HC; "n" varies due to missing data in specific variables.
\*Individual PSS-10, BDI-II, and BAI scores are shown in Supplementary Figure S1.

### Cytokine and inflammatory target assay

Blood samples were obtained by venipuncture, in the general time range 09:00-17:00, at least one hour after the daily MAT dose for the iHUD. Samples were centrifuged (10 minutes at 1,200 G) within ≈1 hour, and serum was stored at -80°C until the time of analysis. Serum samples were analyzed for relative levels of cytokines, using the validated *Olink Target 96 Inflammation* panel (Olink, Uppsala Sweden), following manufacturer’s instructions ^11^, at the Human Immune Monitoring Center of the Icahn School of Medicine at Mount Sinai. This panel measures 92 different targets (principally chemokines, interleukins, growth factors, TNF-related molecules as well as other inflammation-related proteins; full target list: https://olink.com/products-services/target/inflammation/). The assay provides relative quantification of these targets, expressed as normalized protein units (NPX) on a log2 scale. If a specific target in the panel yielded values for which ≥50% of the samples were lower than the limit of detection (LOD) within either the iHUD or HC group, the target was excluded from further analysis ^11^. The remaining targets were analyzed, including individual values <LOD, as in previous studies ^43^. Lastly, individual outliers (>±3SD from the group mean), were removed from the individual target analyses. However, these outliers were later substituted by multiple imputation for the principal component analysis (see below). Of the 92 targets in the assay, 14 were excluded from analysis, because ≥50% of the samples were lower than LOD. These excluded targets were: IL-2RB, IL-1 alpha, IL2, TSLP, IL-22 RA1, Beta-NGF, IL-24, IL13, IL-20, IL33, IL4, LIF, NRTN, and IL5. The remaining 78 targets were therefore analyzed.

### Statistical analyses for demographic and clinical variables

Sex and race distribution across groups was analyzed with Fisher’s exact test. Clinical data (e.g., age, BMI, sleep hours, PSS-10, BDI-II and BAI) were examined across groups with Mann-Whitney U tests.

### Analyses of individual targets

Normalized protein units (NPX) for each target were analyzed with Wilcoxon’s rank-sum tests for group differences, and p-values were corrected for multiple comparisons, using the False Discovery Rate (FDR) approach (5% cutoff level) ^44^.

### Principal component analysis based on 19 cytokines that differed significantly between iHUD and HC

For dimensionality reduction, after identifying 29 targets that showed significant group differences (as above, including correction for multiple comparisons), we focused on the 19 targets among them which are in canonical cytokine families, and conducted a principal component analysis (PCA) ^11,45^. This used centered and z-standardized individual values, and because PCA requires data for all relevant variables, outliers (greater than ±3SD within each group mean) were replaced using a multiple imputation procedure (missMDA in R) ^46^. Nineteen principal continents were calculated in the algorithm, with a 95% threshold for significance based on 1,000 Montecarlo simulations. Differences in principal component scores between iHUD and HC were examined non-parametrically (Mann-Whitney U test). An ROC curve was used to determine if the first principal component (PC1) score could be used as a diagnostic biomarker to separate iHUD and HC groups ^47,48^. For follow-up, Spearman correlations were examined between these PC1 scores and demographic and clinical variables (as in Table 1), with multiple comparison correction (FDR approach; 5% cutoff level).

### Multiple logistic regression with group diagnosis (iHUD vs HC) as binary outcome

PC1 scores based on the 19 cytokines that differed between iHUD and HC (see above) were entered in a multiple logistic regression, together with BMI, age, sex, sleep hours, and PSS-10 (perceived stress) scores. The binary outcome was group diagnosis (HUD vs. HC). An ROC curve was also used to examine the performance of the multiple logistic regression in correctly classifying iHUD vs. HC.

## Results

### Demographics

Table 1 shows demographics and clinical variables. Age and hours slept in the night before testing did not differ between groups. There were relatively more males than females in both iHUD and HC, but the contingency analysis was non-significant. There were relatively more persons of white race in the iHUD versus HC group. Also, iHUD had greater BMI, perceived stress scores (PSS-10), as well as depression (BDI-II), and anxiety (BAI) scores, compared to HC. Supplementary Figure S1 shows that PSS-10 scores are widely distributed across participants, whereas BDI-II and BAI scores showed a robust “floor” effect (i.e., 0 scores), especially in the HC. The mean duration of abstinence in the iHUD was 198 days. Other than MAT in the iHUD, participants had limited usage of other medications (e.g., trazodone for insomnia). None of the participants in either group had current exposure to corticosteroids, other major immunomodulatory or anti-inflammatory medications, or n-acetyl-cysteine.

### Cytokine and inflammatory target data

#### Comparison of iHUD versus HC

The full panel had 92 targets. After excluding 14 targets due to >=50% of the samples being <LOD) (see methods), we compared the remaining 78 targets in iHUD vs. HC with Wilcoxon’s rank-sum tests. After FDR correction, 29 of these targets had significantly different NPX values in iHUD vs. HC. Of these 29 targets (Figure 1A), 26 showed higher levels in iHUD vs. HC. Only 3 targets had the opposite profile (higher in HC vs. iHUD). Data summaries for these 29 targets are in the Supplement Table S1, as well as for the targets that did not reach significance (Supplement Table S2). Figure 1B shows the same data shown as differences in mean scores (i.e., mean iHUD - mean HC), for visualization.

**Figure 1.**
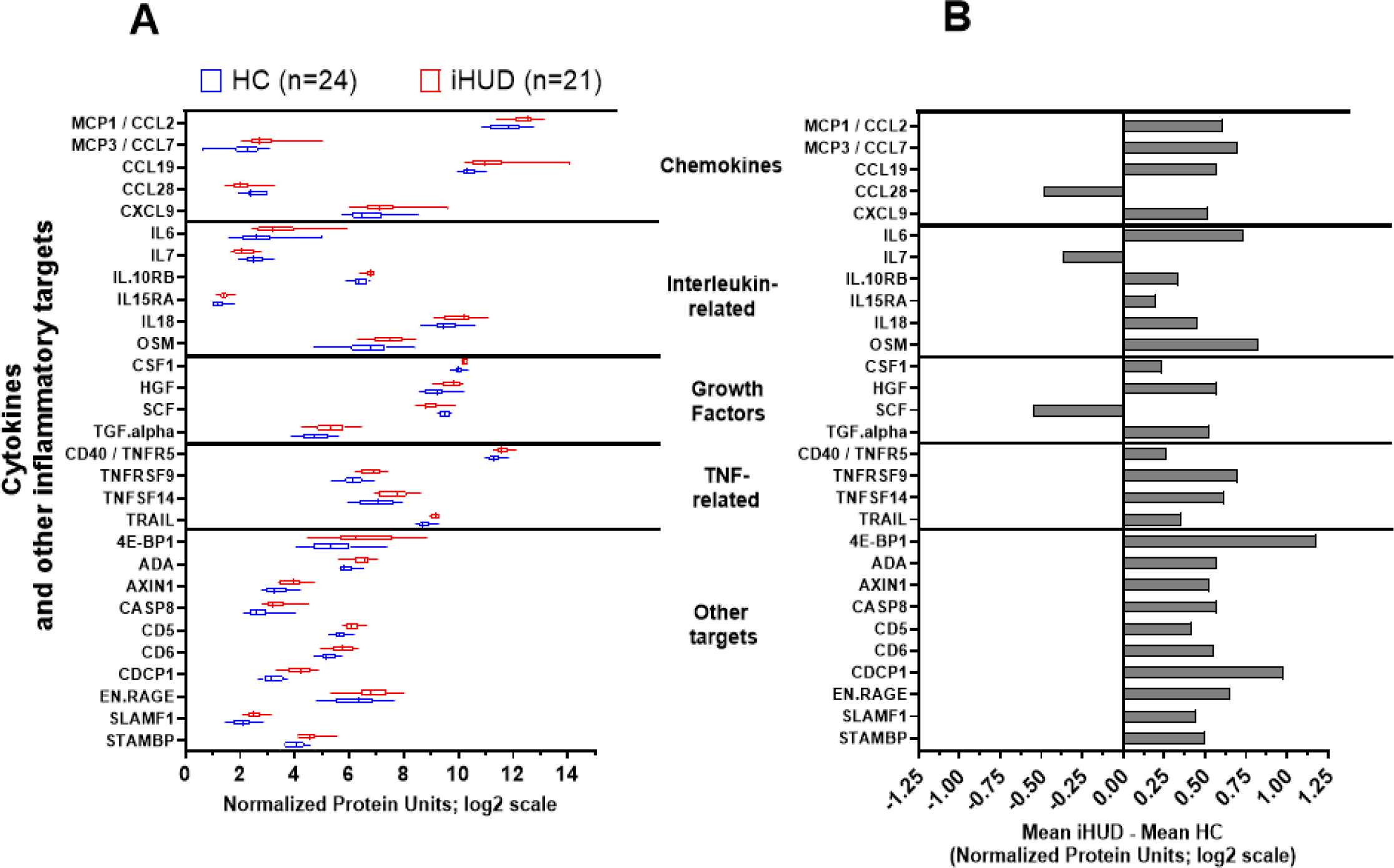
**A.** Box and whisker plot for iHUD and HC, for the 29 targets with significant group differences, after FDR correction. Targets are organized by canonical cytokine families (19 targets; chemokines, interleukin-related, growth factors, and TNF-related), and the remainder as “other targets”. The box marks 25-75 percentiles; midline marks the median and whiskers mark 5-95 percentiles. **B.** For visualization, panel A data are re-plotted as the difference in mean values in iHUD and HC (i.e., mean iHUD - mean HC).

#### Summary of 29 targets with significant differences between iHUD and HC, after multiple comparison correction

Of the 29 targets with significant differences between groups, 19 were members of canonical cytokine families. The cytokines with iHUD>HC levels were the chemokines MCP1/CCL2, MCP3/CCL7, CCL19, and CXCL9, the interleukin-related cytokines were IL6, IL10RB, IL15RA, IL18, OSM, and TRAIL, the growth factors were CSF1, HGF, TGF-alpha and the TNF-related cytokines were TNFRSF9, TNFSF14, and CD40/TNFR5. By contrast, only three cytokines had the opposite profile, with HC>iHUD levels: CCL28, IL7, SCF. All the remaining 10 targets which reached group significance had a iHUD>HC profile. These targets were: eukaryotic translation initiation factor 4E [4E-BP1], adenosine deaminase [ADA], axin-1 (AXIN1], caspase-8 [CASP8], CD5, CD6, CUB domain-containing protein [CDCP1], extracellular newly identified receptor for advanced glycation end-products binding protein [EN.RAGE], signaling lymphocytic activation molecule 1 [SLAMF1] and signal-transducing adaptor molecule-binding protein [STAMBP] (marked “other targets” in Figure 1).

Principal Component Analysis (PCA) on the 19 cytokines that differ significantly in iHUD vs. HC:

Figure 2 shows the PCA results based on the 19 cytokines that differed in iHUD vs. HC (from Fig. 1; excluding 10 significant “other targets” in the assay panel). The Scree plot in Figure 2A shows that the first 2 principal components (i.e., PC1 and PC2) account for 40.9% and 15.8% of variance, respectively (further PC accounted for relatively small proportions of variance). As shown in Figure 2B, PC1 scores were significantly greater in iHUD vs. HC (Mann-Whitney U=36; p<0.0001), whereas PC2 scores did not differ significantly between groups (not shown). A receiver operating characteristic (ROC) of PC1 scores in Figure 2D, with iHUD and HC as the binary outcomes, shows a univariate AUROC=91.7 (p<0.0001). Loadings (Eigenvector*√Eigenvalue) for the 19 cytokines in PC1 are shown in the supplement (Table S3), to illustrate the contribution of individual cytokines to the overall PC score. As expected, the sign of PC loadings differed between the 16 cytokines that had iHUD>HC values versus the 3 cytokines that had HC>iHUD values (i.e., CCL28, IL7 and SCF) (Supplement Table S3).

**Figure 2:**
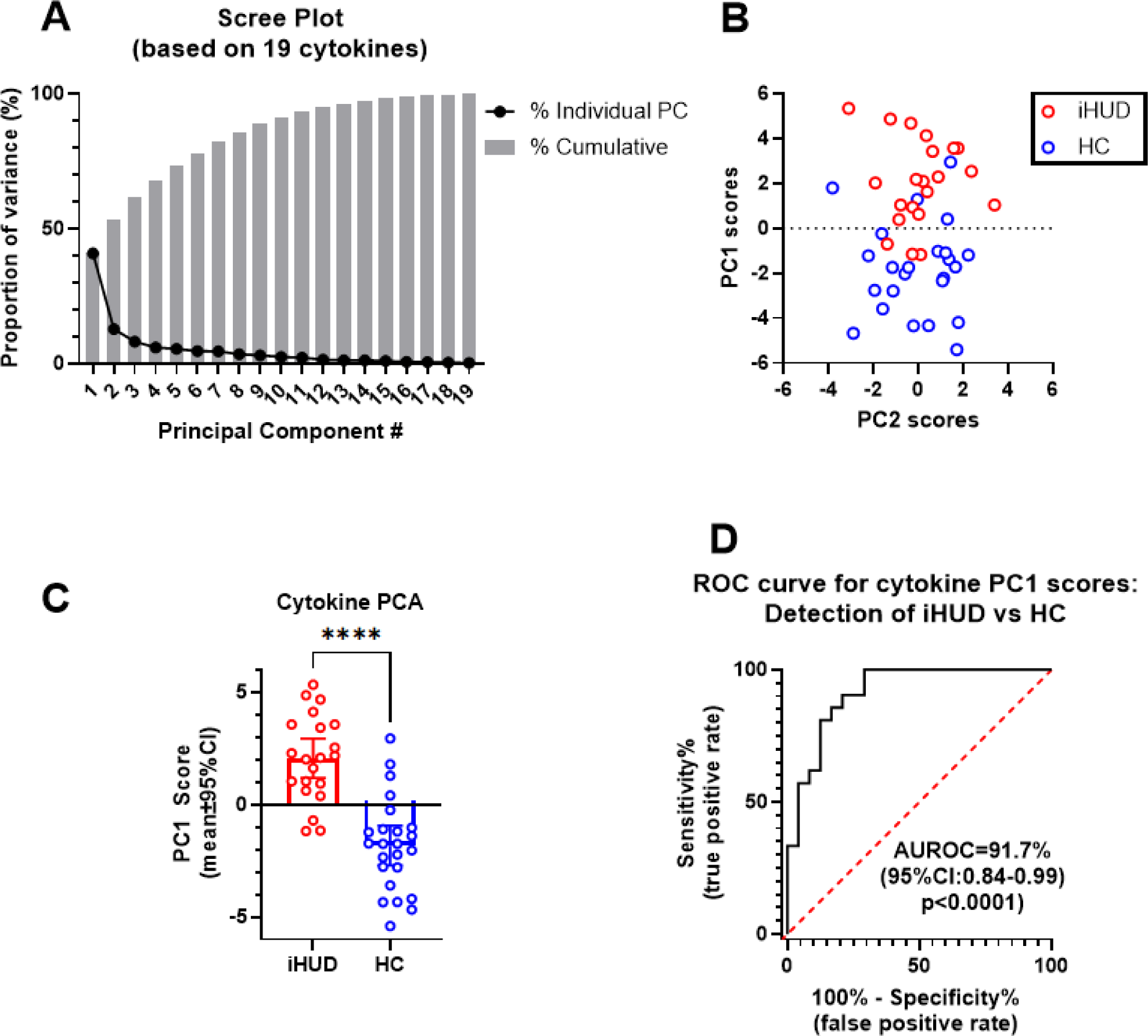
Principal component analysis (PCA) for the 19 cytokines (representing 4 cytokine families in Fig. 1: chemokines, interleukin-related, growth factors, and TNF-related). **A**: Scree plot, showing the proportion of variance accounted for by consecutive principal components (PC). **B:** Scatter plot for individual scores for PC1 and PC2. **C:** Comparison of PC1 scores between iHUD (n=21) and HC (n=24). **D:** Receiver operating characteristic (ROC) curve for PC1 scores as a measure to detect HUD vs HC as binary outcomes.

#### Multiple logistic regression for HUD and HC as binary outcomes, examining cytokine PC1 scores and major demographic and clinical variables

We carried out a multiple logistic regression with group membership as the outcome (iHUD vs. HC), with the following variables: cytokine PC1 scores (from Fig. 2), sex, age, BMI, sleep hours and perceived stress (PSS-10) scores. Depression and anxiety scores (BDI-II and BAI instruments, respectively) were not entered in the regression due to “floor” effects in HC (Supplement Figure S1). Regression parameters are shown in Table 2. In this regression, only cytokine PC1 scores and PSS scores were positive predictors of being in the iHUD category. Overall, the multiple regression had a near unity AUROC (98.3; 95%CI:95.4-100; p<0.0001), indicating excellent effectiveness in differentiating iHUD from HC ^49^.

**Table 2.** Multivariable logistic regression parameters.

| **Variable** | **Odds ratio for**<br>**HUD as outcome** | **(95%CI)** | **p-value** |
| --- | --- | --- | --- |
| **Cytokine PC1 Score\*** | **3.14** | **1.64 to 11.38** | **0.01** |
| **PSS-10**<br>**perceived stress score** | **1.41** | **1.12 to 2.18** | **0.03** |
| **BMI** | 1.47 | 1.04 to 2.76 | 0.09 |
| **AGE** | 0.99 | 0.80 to 1.19 | 0.89 |
| **Sex: Male**<br>**(Female as reference)** | 2.77 | 0.05 to 179.70 | 0.61 |
| **Sleep Hours** | 1.12 | 0.46 to 2.36 | 0.77 |

\*based on 19 cytokines that differ in iHUD vs. HC (see Figs. 1 and 2)

#### Correlations of cytokine PC1 scores with demographic and clinical variables

As shown in the Supplement Table S4, in all participants combined, cytokine PC1 scores were positively correlated with BMI (surviving FDR correction for multiple comparisons). In the iHUD, there were negative correlations of cytokine PC1 scores with BDI-II scores and age of first use of heroin, and a positive correlation with duration of abstinence (none of these survived FDR correction; see Supplement Figure S2).

## Discussion

Using a representative panel of cytokines as well as inflammatory mediators, we found that 29 targets were significantly different between iHUD and HC (26 of these were higher in iHUD vs. HC). Importantly, these 29 targets included 19 members of several major cytokine families (chemokines, interleukins, growth factor and TNF-related), showing robust dysregulation of different cytokine systems in HUD. Several studies, primarily *in vitro*, have shown mechanistic interactions between MOR systems and specific cytokine receptors ^15,50^. Furthermore, chronic exposure to MOR agonists during HUD causes a repeated disruption to the hypothalamic–pituitary–adrenal (HPA) stress axis ^51,52^, and adrenal corticosteroids (e.g., cortisol) have a major modulatory role for diverse cytokines ^53–55^. These are therefore two major types of mechanisms that could underlie robust differences in cytokine levels between iHUD and HC.

### Cytokines with higher levels in iHUD vs. HC

Some of the cytokines that were elevated in iHUD vs HC here, such as the interleukin IL-6, were previously reported ^23,41^. IL-6 has pro-inflammatory effects both peripherally and in the CNS ^56,57^, and also regulates downstream cytokine networks ^17^. Other cytokines that were elevated in iHUD vs HC, including the chemokine CCL2 (ligand for CCR2), were recently shown to mediate neuro-glial adaptations after MOR-agonist exposure ^18^. Furthermore, genes for several of the cytokines that were elevated in iHUD versus HC (or their cognate receptors) exhibited changes in regional brain expression in rodents exposed to MOR agonists, including CCL2, hepatocyte growth factor (HGF), oncostatin M (OSM) and colony-stimulating factor-1 (CSF1) ^58–60^. Overall, the specific functions and status of these targets as markers of disease severity in iHUD are important areas for future study.

### Cytokines with lower levels in iHUD vs. HC

Only three of the cytokines in the assay were significantly lower in iHUD vs. HC, after multiple comparison correction: stem cell factor (SCF), interleukin IL-7, and the chemokine CCL28. A small number of studies have reported interactions between these cytokines and opioid receptor systems, either *in vitro* or in animal models ^61–63^. However, to our knowledge, this is the first report to show differences in their serum levels in iHUD vs. HC. Therefore future studies should examine their potential mechanistic relevance to this disorder.

### Other inflammatory targets (not part of canonical cytokine families)

Ten proteins that are not part of canonical cytokine families had higher levels in iHUD vs. HC (see Figure 1). Functional changes in some of those targets have been observed after MOR-agonist exposure (especially ADA, adenosine deaminase, and CASP8, caspase-8] ^64,65^. Future studies could determine the functional correlates of these targets, in iHUD vs. HC. To our knowledge, some other targets that differed significantly in iHUD versus HC have not been previously associated with MOR-agonist exposure either *in vitro* or *in vivo*. The mechanistic underpinning of changes in these targets in HUD should also be determined.

### Developing a “cytokine biomarker score” to differentiate iHUD from HC

In addition to documenting differences in specific serum cytokine and inflammatory proteins of iHUD vs. HC, this study also identified a robust blood-based biomarker score (based on PC1 scores from 19 cytokines that differed in iHUD vs. HC, as discussed above) that is of potential diagnostic value as a positive predictor of being in the HUD class, since the univariate AUROC of 91.7%, considered in the “excellent” range ^49^. The diagnostic value of this PC1 cytokine biomarker score survived adjustment for major variables known to affect cytokine markers (e.g., age, sex, BMI and sleep) in a multiple logistic regression ^29,30,66,67^. In this multiple regression, the perceived stress score was also a positive predictor of being in the iHUD category. This is consistent with the role of stress exposure in the severity of HUD, and in dysregulation of the HPA-stress axis (which can itself affect cytokine systems) ^21,68^. In follow-ups, this cytokine PC1 score was correlated with specific aspects of age trajectory in the iHUD (although this did not survive FDR correction). Specifically, in the iHUD, the cytokine PC1 score was negatively correlated with depression BDI-II scores and with age of onset of regular heroin use. Depression signs and age trajectory of heroin are important facets of HUD history and severity ^27,39^. Intriguingly, cytokine PC1 scores were positively correlated with duration of heroin abstinence. However, for all these correlations, larger samples are necessary to determine linearity of these relationships with cytokine PC1 scores (and with specific cytokines therein), while adjusting for demographic features.

### Methodological considerations

These iHUD were receiving standard-of-care MAT (primarily methadone). However, the daily methadone dose was not correlated with cytokine PC1 scores, suggesting that the robust difference in cytokine PC1 scores between iHUD and HC are unlikely to be primarily driven by methadone *per se*. In a follow-up, we also examined the performance of the above multiple logistic regression, using only the n=17 with methadone maintenance (i.e. excluding n=4 with buprenorphine). In this subset analysis, cytokine PC1 scores remained a positive predictor of HUD, adjusting for the other variables (not shown). More broadly, we employed a simplified two-step approach for dimensionality reduction: first focusing on 29 targets that differed significantly between iHUD and HC, and secondly with PCA based on a subset of 19 targets that are members of canonical cytokine families ^17,23^. Other methods to elucidate the optimal components of a “cytokine PC score” for use in iHUD can also be evaluated in larger studies. As a separate issue, while circadian effects have been detected for specific cytokines (e.g., IL6) ^69^, the present samples were obtained across a relatively broad range in daytime hours. However, this time range did not vary systematically across iHUD and HC, therefore, it is unlikely that the robust group differences herein are mainly due to circadian effects.

### Future directions

Prior studies have used cytokine levels as biomarkers of other neuropsychiatric conditions such as anxiety and depression/anhedonia [7,50–52]. Some recent studies have shown that differences in levels of specific cytokines can normalize over prolonged opioid abstinence, (e.g., for IL6) [18]. Therefore, future studies in larger cohorts should determine if this multi-target cytokine biomarker score also differs across stages in HUD recovery trajectory. Another crucial avenue for future research involves investigating sex differences ^1^. Studies have indicated that women with some substance use disorders exhibit heightened susceptibility to stress exposure, and this profile could potentially result in sexually dimorphic cytokine dysregulation ^21,66^; however sex differences in cytokine responses to MOR agonists have not been investigated in depth in humans ^21^.

### Conclusions and future studies

This is one of the few studies to examine a comprehensive set of cytokines from several major families, and detected robust differences in levels of both previously known and novel targets ^21^, in iHUD compared to HC. Cytokines are known to act in interactive networks, both in the periphery and brain ^17^, therefore, it is important to consider cytokine effects as a group, and not only as individual targets. In this regard, this is also the first study to provide a multi-target “cytokine biomarker score” ^42,70^ that was a positive predictor of being in the iHUD group, surviving adjustment for major demographic and clinical variables. Since perceived stress scores were also positive predictors in the multivariable model, future larger-scale studies could determine whether stress mechanisms are directly related to the cytokine differences observed between groups ^28^. It is also important to determine whether such a multi-target cytokine biomarker score (or an optimized version thereof) can be applied to individuals at different stages in the trajectory of HUD and recovery, and whether it is related to the severity of use, abstinence, or relapse ^71^.

## Supporting information

Supplementary Results

## Data Availability

Data produced in the present study are available upon reasonable request to the authors, subject to IRB, consent and privacy issues.

## Abbreviations

AUROC: Area under the receiver-operating curve
BMI: Body mass index
FDR: False discovery rate
HGF: Hepatocyte growth factor
HC: Healthy controls
HUD: Heroin use disorder
iHUD: Individuals with heroin use disorder
IL: interleukin
IQR: Inter-quartile range
LOD: limit of detection
MOR: Mu-opioid receptors
NPX: Normalized protein expression (relative quantification from Olink assay; log2 units)
OSM: Oncostatin M
PC1 scores: Principal component 1 scores
PCA: Principal component analysis
ROC: Receiver-operating curve
SCF: Stem cell factor
TNF: Tumor necrosis factor

## Acknowledgments

We are very grateful to all the clinical coordinators of the NARC laboratory.

## Author Contributions

Study conception and design: (RZG, NAK, YH, ERB); data collection and sample preparation (YH, PR, FC); analysis and interpretation of results (FC, SJR, YH, ERB, NAK, RZG); draft manuscript preparation: (ERB, YH, NAK, RZG, POG). All authors reviewed the results and approved the final version of the manuscript.

