## Supplementary Results for "Serum cytokine and inflammatory markers in individuals with heroin use disorder: potential biomarkers for diagnosis and disease severity"

**Supplement Figure S1:** Individual Beck Depression Inventory-II (BDI-II), Beck Anxiety Inventory (BAI) and Perceived Stress Scale (PSS-10) scores, in iHUD and HC in panels A-C, respectively; with line indicating median. Data were analyzed with Mann-Whitney U tests, as shown in Table 1. Note the frequent “0” (i.e., floor) scores especially in healthy controls, for BDI-II and BAI scales. iHUD “n” differs due to missing data.

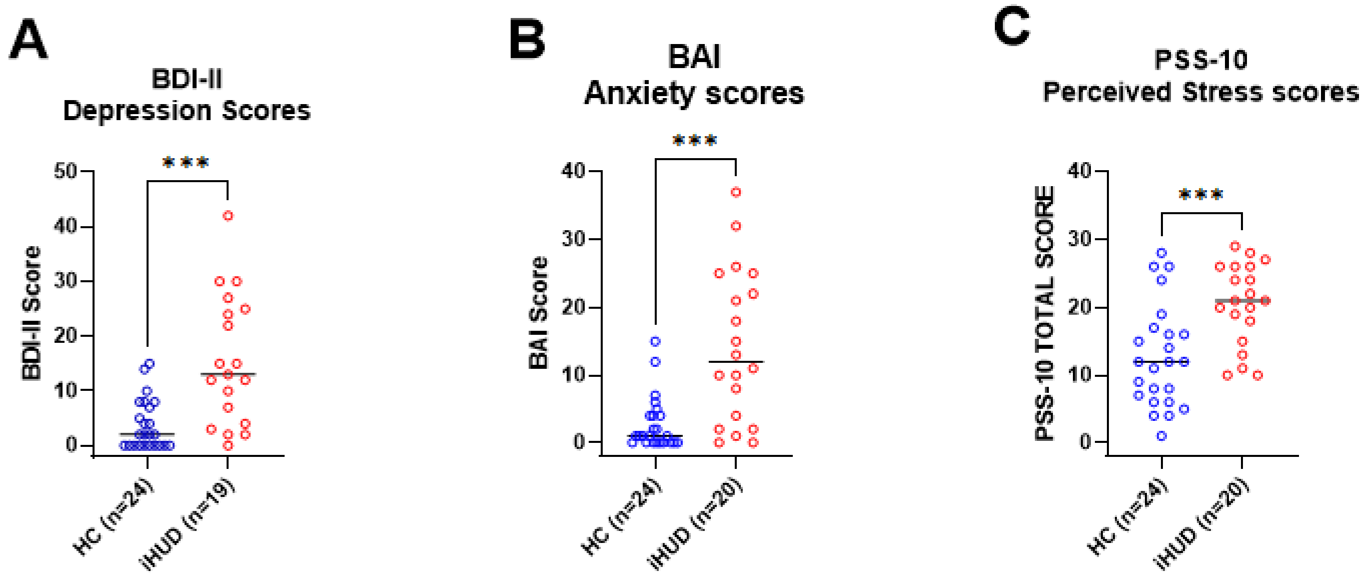

**Table S1. Data for the 29 targets with significant differences in iHUD versus HC after False Discovery Rate (FDR) correction. Targets in bold italics indicate the 19 cytokines from canonical families that are used in PCA analysis**

| Target | Family | Used in PCA? | iHUD (n=21) | | | HC (n=24) | | | iHUD vs HC comparison | | |
|---|---|---|---|---|---|---|---|---|---|---|---|
| | | | Mean NPX units, log2 scale | 95%CI | Outliers Replaced | Mean NPX units, log2 scale | 95%CI | Outliers Replaced | W statistic | Uncorrected p value | FDR p value |
| ***MCP1 / CCL2*** | Chemokines | Yes | 12.38 | 0.24 | 0 | 11.77 | 0.28 | 0 | 123 | 0.0028 | 0.0116 |
| ***MCP3 / CCL7*** | Chemokines | Yes | 2.91 | 0.36 | 0 | 2.21 | 0.26 | 0 | 122 | 0.0026 | 0.0113 |
| ***CCL19*** | Chemokines | Yes | 10.97 | 0.31 | 1 | 10.40 | 0.16 | 1 | 112 | 0.0035 | 0.0130 |
| ***CCL28*** | Chemokines | Yes | 2.06 | 0.21 | 0 | 2.54 | 0.17 | 0 | 408 | 0.0002 | 0.0019 |
| ***CXCL9*** | Chemokines | Yes | 7.10 | 0.27 | 1 | 6.58 | 0.31 | 1 | 118 | 0.0057 | 0.0195 |
| ***IL6*** | Interleukin | Yes | 3.44 | 0.46 | 1 | 2.70 | 0.37 | 0 | 126 | 0.0066 | 0.0205 |
| ***IL7*** | Interleukin | Yes | 2.15 | 0.21 | 0 | 2.52 | 0.18 | 0 | 362 | 0.0117 | 0.0326 |
| ***IL10RB*** | Interleukin | Yes | 6.75 | 0.10 | 0 | 6.40 | 0.13 | 0 | 98 | 0.0003 | 0.0021 |
| ***IL15RA*** | Interleukin | Yes | 1.41 | 0.11 | 0 | 1.21 | 0.11 | 0 | 132 | 0.0057 | 0.0195 |
| ***IL18*** | Interleukin | Yes | 10.02 | 0.28 | 0 | 9.56 | 0.24 | 0 | 137 | 0.0083 | 0.0248 |
| ***OSM*** | Interleukin | Yes | 7.47 | 0.33 | 0 | 6.64 | 0.40 | 0 | 122 | 0.0026 | 0.0113 |
| ***CSF1*** | Growth factor | Yes | 10.22 | 0.07 | 0 | 9.98 | 0.10 | 0 | 93 | 0.0002 | 0.0017 |
| ***HGF*** | Growth factor | Yes | 9.71 | 0.20 | 0 | 9.14 | 0.22 | 0 | 93 | 0.0002 | 0.0017 |
| ***SCF*** | Growth factor | Yes | 8.95 | 0.20 | 0 | 9.50 | 0.11 | 0 | 431 | $1.7X10^{-5}$ | 0.0004 |
| ***TGF.alpha*** | Growth factor | Yes | 5.30 | 0.29 | 0 | 4.77 | 0.24 | 0 | 133 | 0.0062 | 0.0200 |
| ***CD40 / TNFR5*** | TNF-related | Yes | 11.61 | 0.14 | 0 | 11.35 | 0.13 | 0 | 124 | 0.0031 | 0.0120 |
| ***TNFRSF9*** | TNF-related | Yes | 6.80 | 0.22 | 0 | 6.10 | 0.17 | 1 | 66 | $1.23X10^{-5}$ | 0.0004 |
| ***TNFSF14*** | TNF-related | Yes | 7.63 | 0.29 | 0 | 7.01 | 0.30 | 0 | 140 | 0.0102 | 0.0295 |
| ***TRAIL*** | TNF-related | Yes | 9.15 | 0.09 | 0 | 8.79 | 0.13 | 0 | 89 | 0.0001 | 0.0015 |
| 4E-BP1 | Other | No | 6.57 | 0.55 | 0 | 5.39 | 0.42 | 1 | 108 | 0.0013 | 0.0065 |
| ADA | Other | No | 6.46 | 0.20 | 0 | 5.89 | 0.15 | 1 | 76 | $4.49X10^{-5}$ | 0.0007 |
| AXIN1 | Other | No | 3.88 | 0.21 | 0 | 3.35 | 0.22 | 0 | 114 | 0.0013 | 0.0065 |
| CASP8 | Other | No | 3.30 | 0.23 | 0 | 2.73 | 0.24 | 0 | 103 | 0.0005 | 0.0029 |

| | | | | | | | | | | | |
|---|---|---|---|---|---|---|---|---|---|---|---|
| CD5 | Other | No | 6.12 | 0.15 | 0 | 5.70 | 0.12 | 0 | 76 | $2.48X10^{-5}$ | 0.0005 |
| CD6 | Other | No | 5.76 | 0.23 | 0 | 5.20 | 0.15 | 1 | 90 | 0.0002 | 0.0019 |
| CDCP1 | Other | No | 4.18 | 0.24 | 1 | 3.20 | 0.18 | 0 | 34 | $7.35X10^{-8}$ | $5.73X10^{-6}$ |
| EN.RAGE | Other | No | 6.84 | 0.34 | 0 | 6.18 | 0.37 | 0 | 145 | 0.0143 | 0.0386 |
| SLAMF1 | Other | No | 2.51 | 0.16 | 0 | 2.06 | 0.18 | 0 | 103 | 0.0005 | 0.0029 |
| STAMBP | Other | No | 4.54 | 0.21 | 0 | 4.04 | 0.17 | 1 | 99 | 0.0006 | 0.0032 |

Table S2. Data for 49 targets ***without*** significant differences between iHUD and HC after FDR correction

| Target | iHUD (n=21) | | HC (n=24) | | iHUD vs HC comparison | | |
|---|---|---|---|---|---|---|---|
| | Mean NPX units, log2 scale | 95%CI | Mean NPX units, log2 scale | 95%CI | W statistic | Unadjusted p value | FDR p value |
| **uPA** | 10.71 | 0.11 | 10.51 | 0.14 | 151 | 0.02 | 0.06 |
| **IL.18R1** | 7.27 | 0.16 | 6.98 | 0.15 | 153 | 0.02 | 0.06 |
| **TNFB** | 5.01 | 0.22 | 4.75 | 0.18 | 158 | 0.03 | 0.08 |
| **CXCL1** | 9.11 | 0.29 | 9.77 | 0.41 | 344 | 0.04 | 0.08 |
| **IL10** | 3.34 | 0.23 | 3.00 | 0.23 | 160 | 0.04 | 0.08 |
| **TNF** | 4.00 | 0.15 | 3.84 | 0.22 | 153 | 0.04 | 0.08 |
| **SIRT2** | 2.79 | 0.29 | 2.33 | 0.40 | 161 | 0.04 | 0.08 |
| **IL.17C** | 2.20 | 0.20 | 2.67 | 0.35 | 325 | 0.05 | 0.10 |
| **TRANCE** | 5.59 | 0.30 | 5.17 | 0.31 | 167 | 0.05 | 0.11 |
| **Flt3L** | 9.85 | 0.20 | 9.58 | 0.18 | 169 | 0.06 | 0.12 |
| **CXCL6** | 9.18 | 0.32 | 9.79 | 0.39 | 335 | 0.06 | 0.12 |
| **IFN.gamma** | 7.46 | 0.36 | 7.01 | 0.34 | 170 | 0.06 | 0.12 |
| **MMP.1** | 15.44 | 0.26 | 15.80 | 0.33 | 333 | 0.07 | 0.12 |
| **CD8A** | 10.30 | 0.30 | 9.90 | 0.31 | 177 | 0.09 | 0.16 |
| **IL.12B** | 6.87 | 0.32 | 6.53 | 0.31 | 177 | 0.09 | 0.16 |
| **CCL23** | 11.74 | 0.20 | 11.48 | 0.20 | 177 | 0.09 | 0.16 |
| **CD244** | 5.55 | 0.15 | 5.39 | 0.15 | 180 | 0.10 | 0.18 |
| **MMP.10** | 8.91 | 0.28 | 8.65 | 0.20 | 182 | 0.11 | 0.19 |
| **VEGFA** | 12.90 | 0.23 | 12.65 | 0.21 | 184 | 0.13 | 0.20 |
| **MCP.4** | 15.72 | 0.23 | 15.91 | 0.21 | 314 | 0.16 | 0.26 |
| **CXCL10** | 8.37 | 0.39 | 8.04 | 0.34 | 191 | 0.17 | 0.27 |
| **GDNF** | 1.72 | 0.14 | 1.87 | 0.15 | 300 | 0.17 | 0.27 |
| **CCL25** | 6.32 | 0.25 | 6.64 | 0.34 | 311 | 0.19 | 0.28 |
| **DNER** | 9.39 | 0.10 | 9.48 | 0.12 | 308 | 0.21 | 0.31 |
| **FGF.23** | 0.85 | 0.25 | 0.73 | 0.24 | 190 | 0.23 | 0.34 |
| **LAP.TGF.beta.1** | 7.13 | 0.10 | 7.22 | 0.13 | 299 | 0.29 | 0.41 |

| | | | | | | | |
|---|---|---|---|---|---|---|---|
| **TWEAK** | 10.00 | 0.16 | 9.90 | 0.14 | 205 | 0.29 | 0.41 |
| **CXCL5** | 12.56 | 0.39 | 12.91 | 0.34 | 298 | 0.30 | 0.42 |
| **CCL11** | 8.20 | 0.19 | 8.16 | 0.30 | 208 | 0.33 | 0.44 |
| **LIF.R** | 3.57 | 0.11 | 3.66 | 0.08 | 292 | 0.37 | 0.49 |
| **IL.20RA** | 0.41 | 0.16 | 0.53 | 0.22 | 277 | 0.39 | 0.51 |
| **PD.L1** | 5.75 | 0.16 | 5.71 | 0.18 | 204 | 0.41 | 0.52 |
| **CX3CL1** | 4.60 | 0.18 | 4.69 | 0.19 | 285 | 0.46 | 0.58 |
| **FGF.19** | 8.93 | 0.24 | 8.86 | 0.33 | 226 | 0.57 | 0.70 |
| **IL.10RA** | 1.13 | 0.31 | 1.07 | 0.29 | 230 | 0.63 | 0.77 |
| **OPG** | 9.92 | 0.16 | 9.91 | 0.13 | 232 | 0.66 | 0.78 |
| **ST1A1** | 4.67 | 0.53 | 4.56 | 0.47 | 232 | 0.66 | 0.78 |
| **CXCL11** | 8.28 | 0.50 | 8.55 | 0.56 | 271 | 0.68 | 0.79 |
| **ARTN** | 0.19 | 0.13 | 0.16 | 0.09 | 224 | 0.72 | 0.82 |
| **MCP.2** | 9.80 | 0.27 | 9.91 | 0.24 | 268 | 0.73 | 0.82 |
| **NT.3** | 2.58 | 0.20 | 2.54 | 0.13 | 238 | 0.76 | 0.85 |
| **CCL4** | 7.62 | 0.26 | 7.57 | 0.29 | 247 | 0.92 | 0.97 |
| **FGF.5** | 1.66 | 0.08 | 1.69 | 0.14 | 257 | 0.92 | 0.97 |
| **FGF.21** | 4.95 | 0.69 | 4.89 | 0.59 | 257 | 0.92 | 0.97 |
| **CCL20** | 7.58 | 0.26 | 7.69 | 0.41 | 247 | 0.92 | 0.97 |
| **IL8** | 6.98 | 0.43 | 6.93 | 0.39 | 226 | 0.93 | 0.97 |
| **IL.17A** | 0.68 | 0.17 | 0.74 | 0.22 | 239 | 0.96 | 0.98 |
| **CCL3** | 6.98 | 0.31 | 7.01 | 0.37 | 239 | 0.96 | 0.98 |
| **CST5** | 7.70 | 0.24 | 7.76 | 0.28 | 252 | 1.00 | 1.00 |

**Table S3. PC1 Loadings for 19 cytokine components of PCA**

| Cytokine | Family | PC1 Loading (Eigenvector*√Eigenvalue) |
|---|---|---|
| **MCP.1 / CCL2** | Chemokines | 0.55 |
| **MCP.3 / CCL7** | Chemokines | 0.71 |
| **CCL19** | Chemokines | 0.61 |
| **CCL28** | Chemokines | -0.19 |
| **CXCL9** | Chemokines | 0.67 |
| **IL7** | Interleukin | -0.22 |
| **IL6** | Interleukin | 0.55 |
| **IL.10RB** | Interleukin | 0.59 |
| **IL.15RA** | Interleukin | 0.69 |
| **IL18** | Interleukin | 0.63 |
| **OSM** | Interleukin | 0.70 |
| **CSF.1** | Growth factor | 0.86 |
| **HGF** | Growth factor | 0.79 |
| **SCF** | Growth factor | -0.55 |
| **TGF.alpha** | Growth factor | 0.60 |
| **CD40 /TNFR5** | TNF-related | 0.67 |

| | | |
|---|---|---|
| **TNFRSF9** | TNF-related | 0.77 |
| **TNFSF14** | TNF-related | 0.68 |
| **TRAIL** | TNF-related | 0.72 |

**Table S4. Correlation of cytokine PC1 scores (19 cytokines, see tables S1 and S3) with demographic and clinical variables**

| Variable | Spearman correlations of cytokine PC1 scores with clinical and phenotypic variables | | | | | |
|---|---|---|---|---|---|---|
| | *iHUD only (n=21 unless otherwise stated)* | | *HC only (n=24 unless otherwise stated)* | | *iHUD and HC (n=45 unless otherwise stated)* | |
| | **R (uncorrected p)** | **FDR (uncorrected p)** | **R (uncorrected p)** | **FDR (corrected p)** | **R (uncorrected p)** | **FDR (corrected p)** |
| **Age** | -0.21 (0.36) | NS | 0.14 (0.52) | NS | 0.084 (0.59) | NS |
| **BMI** | 0.0078 (0.97) | NS | 0.34 (0.11) | NS | **0.43 (0.0031)** | **p=0.012** |
| **Sleep hours in the prior night** | 0.046 (0.85) | NS | 0.33 (0.11) | NS | 0.19 (0.22) | NS |
| **Perceived Stress PSS-10 scores** | -0.35 (0.14) n=20 | NS | -0.32 (0.13) | NS | 0.16 (0.3) n=44 | NS |
| **Depression BDI-II scores** | **-0.49 (0.032) n=19** | NS | Not calculated due to “floor” effect in HC; See Supplement Fig. S1 | | | |
| **Anxiety BAI scores** | -0.12 (0.60) n=20 | NS | | | | |
| **Age of onset of first use of heroin** | -0.42 (0.069) | NS | N/A | N/A | N/A | N/A |
| **Age of onset of regular use of heroin** | **-0.48 (0.031)** | NS | N/A | N/A | N/A | N/A |
| **Years of regular heroin use** | -0.23 (0.35) | NS | N/A | N/A | N/A | N/A |
| **Heroin** | **0.50 (0.02)** | NS | N/A | N/A | N/A | N/A |

| | | | | | | |
|---|---|---|---|---|---|---|
| **abstinence duration** | | | | | | |
| **Methadone daily dose in mg** | 0.25 (0.34) n=16 | NS | N/A | N/A | N/A | N/A |

**FDR: False discovery rate; 5% cutoff level**

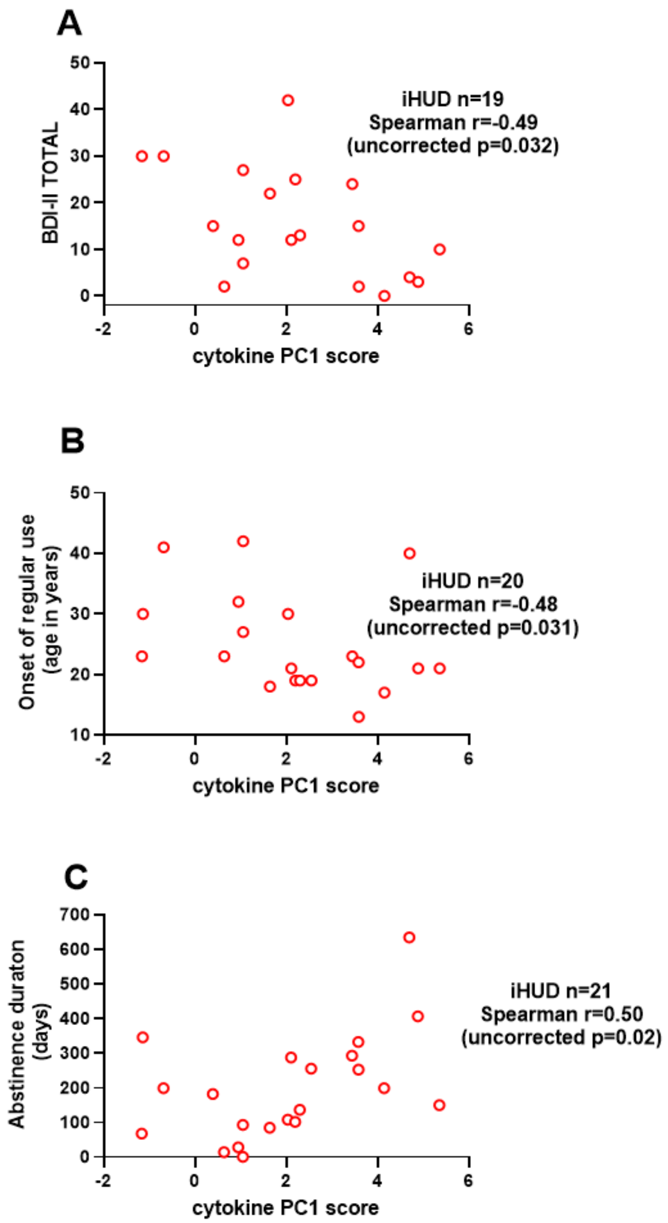


**Figure S2:** Correlation of individual cytokine PC1 scores with BDI-II scores, age of onset of regular heroin use and abstinence duration in iHUD (panels A,B,C respectively); “n” differs across panels due to missing data.
